## Appendices for "COVID-19 vaccine hesitancy and resistance: Correlates in a nationally representative longitudinal survey of the Australian population"

### 1 Appendix

#### 1.1 Covariates

Demographic variables included sex, age (18-24, 25-34, 35-44, 45-54, 55-64, 65-74 and 75 years or more), indigenous status, education (less than Year 12, Year 12, above Year 12 but below a bachelor’s degree and bachelor’s degree or above), born overseas (English speak or non-English speaking country), speaks a language other than English at home, capital or non-capital city, and employment status (employed, not employed). Continuous household income was estimated based on an interval regression model using responses to a categorical income question (with ten possible income categories). Socio-economic status of the area: Socio-economic indexes for areas (SEIFA) quintiles from most disadvantaged to most advantage. SEIFA was measured using the Index of Relative Social Advantage or Disadvantage (ABS, 2016).

Health related variables included self-rated health and disability or chronic illness. Self-rated health was measured using “How is your health in general? Would you say it is…?” with participants reporting very good, good, fair, bad or very bad. Disability or chronic illness was measured using an item from the European Social Survey “Are you hampered in your daily activities in any way by any longstanding illness, or disability, infirmity, or mental health problem?” Participants reported “Yes, a lot”, “Yes, to some extent” or “No”.

##### 1.1.1 COVID-19 related variables.

Social distancing: Participants were asked whether they did any of the following in the past week: avoid crowded places, avoid public places, keep your distance from others (1.5 metres), quarantine yourself if you have symptoms, wore a face mask indoors when in a public place, and wore a mask outdoors when in a public place. These variables were highly correlated with each other, and were combined using principal components analysis to create a COVID-19 behaviour index that has been scaled to have mean of zero and standard deviation of one.

Tested for COVID-19 was measured by the whether participants reported that they had been tested by a doctor or nurse for COVID-19.

Concerns about COVID-19 was measured by “You felt anxious or worried for the safety of yourself, close family members or friends, due to COVID-19”.

Scepticism about the COVID-19 was captured by asking participants the extent to which they agreed with the following statement “There has been too much unnecessary worry about the COVID-19 outbreak” (strongly agree to strongly disagree).

COVIDSafe app. The Australian Federal Government developed the COVIDSafe app to help state and territory health officials to quickly contact people who may have been exposed to COVID-19. However, some people have expressed concern about the security of the COVIDSafe app and whether it will be used for mass surveillance so therefore we asked a question about the whether participants had downloaded the app. We asked “Have you installed the COVIDSafe app on your phone?”, participants could respond yes or no.

##### 1.1.2 Political, and social attitudes.

Voting intentions*.* Participants were asked “If a federal election for the House of Representatives was held today, which one of the following parties would you vote for?” they could indicate they would vote for the conservative coalition (Liberal Party and National Party), the Labour party, the Greens, some other party or they did not know.

Religiosity was measured using the following item from the European Social Survey “Regardless of whether you belong to a particular religion, how religious would you say you are?” on a scale of 0 (Not at all) to 10 (Very) asked in February 2020.

Support for migration was measured by asking “Is Australia made a worse or a better place to live by people coming to live here from other countries?” on a scale of 0 (Not at all) to 10 (Very). Participants were asked this item in February 2020 and is taken from the European Social Survey.

Social trust was measured using the following questions “Generally speaking, would you say that most people can be trusted, or that you can’t be too careful in dealing with people?”, “Do you think that most people would try to take advantage of you if they got the chance, or would they try to be fair?”, and “Would you say that most of the time people try to be helpful or that they are mostly looking out for themselves?”. For each item participants were asked to rate these items on a scale of 0 to 10 and for our analyses we averaged ratings on these three items.

Confidence in the Federal, state government and hospitals and the health system was rated by each participant (1: a great deal, 2: quite a lot, 3: not very much, 4: none at all).

Populism was measured using items from the June 2019 Comparative Study of Electoral Systems (CSES): “What people call compromise in politics is really just selling out on one's principles”, “Most politicians do not care about the people”, “Politicians are the main problem in Australia”, “The people, and not politicians, should make our most important policy decisions”, and “Most politicians care only about the interests of the rich and powerful” (reversed). Respondents were given five options: 1. Strongly agree; 2. Somewhat agree; 3. Neither agree nor disagree; 4. Somewhat disagree; and 5. Strongly disagree and items were added to form an index.

Authoritarianism was asked in February 2020 using the following item from the European Social Survey: “Having a strong leader in government is good for Australia even if the leader bends the rules to get things done”. Participants were asked to rate strongly agree, somewhat agree, neither agree nor disagree, somewhat disagree, or strongly disagree.

Altruism is based on responses to a question in February 2020 as to whether the individual thought the following description matched themselves: ‘It's very important to him/her to help the people around him/her. He/She wants to care for their well-being’

**Table A.1	Descriptive statistics – Model 1**

| Variable | Mean | SD | Minimum | Maximum |
| --- | --- | --- | --- | --- |
| Vaccination intentions | 3.40 | 0.85 | 1.00 | 4.00 |
| Victoria | 0.26 | 0.44 | 0.00 | 1.00 |
| Female | 0.50 | 0.50 | 0.00 | 1.00 |
| Aged 18 to 24 years | 0.08 | 0.27 | 0.00 | 1.00 |
| Aged 25 to 34 years | 0.22 | 0.41 | 0.00 | 1.00 |
| Aged 45 to 54 years | 0.17 | 0.38 | 0.00 | 1.00 |
| Aged 55 to 64 years | 0.15 | 0.36 | 0.00 | 1.00 |
| Aged 65 to 74 years | 0.13 | 0.34 | 0.00 | 1.00 |
| Aged 75 years plus | 0.06 | 0.24 | 0.00 | 1.00 |
| Indigenous | 0.02 | 0.14 | 0.00 | 1.00 |
| Born overseas in a main English speaking country | 0.11 | 0.31 | 0.00 | 1.00 |
| Born overseas in a non-English speaking country | 0.23 | 0.42 | 0.00 | 1.00 |
| Speaks a language other than English at home | 0.24 | 0.42 | 0.00 | 1.00 |
| Has not completed Year 12 or post-school qualification | 0.14 | 0.35 | 0.00 | 1.00 |
| Has a post graduate degree | 0.09 | 0.28 | 0.00 | 1.00 |
| Has an undergraduate degree | 0.17 | 0.38 | 0.00 | 1.00 |
| Has a Certificate III/IV, Diploma or Associate Degree | 0.40 | 0.49 | 0.00 | 1.00 |
| Lives in the most disadvantaged areas (1st quintile) | 0.18 | 0.39 | 0.00 | 1.00 |
| Lives in next most disadvantaged areas (2nd quintile) | 0.20 | 0.40 | 0.00 | 1.00 |
| Lives in next most advantaged areas (4th quintile) | 0.21 | 0.40 | 0.00 | 1.00 |
| Lives in the most advantaged areas (5th quintile) | 0.21 | 0.41 | 0.00 | 1.00 |
| Lives in a non-capital city | 0.33 | 0.47 | 0.00 | 1.00 |
| Employed | 0.60 | 0.49 | 0.00 | 1.00 |
| Household income | 670.81 | 541.96 | -8.86 | 6421.05 |

Source:	ANUpoll, April, May and August 2020.

Notes:	Descriptive statistics from Ordered probit model. N = 2,717.

**Table A.2	Descriptive statistics – Final model**

| Variable | Mean | SD | Minimum | Maximum |
| --- | --- | --- | --- | --- |
| Vaccination intentions | 3.40 | 0.86 | 1.00 | 4.00 |
| Victoria | 0.25 | 0.44 | 0.00 | 1.00 |
| Female | 0.49 | 0.50 | 0.00 | 1.00 |
| Aged 18 to 24 years | 0.08 | 0.27 | 0.00 | 1.00 |
| Aged 25 to 34 years | 0.21 | 0.41 | 0.00 | 1.00 |
| Aged 45 to 54 years | 0.17 | 0.38 | 0.00 | 1.00 |
| Aged 55 to 64 years | 0.16 | 0.36 | 0.00 | 1.00 |
| Aged 65 to 74 years | 0.13 | 0.34 | 0.00 | 1.00 |
| Aged 75 years plus | 0.06 | 0.24 | 0.00 | 1.00 |
| Indigenous | 0.02 | 0.13 | 0.00 | 1.00 |
| Born overseas in a main English speaking country | 0.11 | 0.31 | 0.00 | 1.00 |
| Born overseas in a non-English speaking country | 0.24 | 0.42 | 0.00 | 1.00 |
| Speaks a language other than English at home | 0.24 | 0.43 | 0.00 | 1.00 |
| Has not completed Year 12 or post-school qualification | 0.14 | 0.34 | 0.00 | 1.00 |
| Has a post graduate degree | 0.08 | 0.28 | 0.00 | 1.00 |
| Has an undergraduate degree | 0.17 | 0.38 | 0.00 | 1.00 |
| Has a Certificate III/IV, Diploma or Associate Degree | 0.40 | 0.49 | 0.00 | 1.00 |
| Lives in the most disadvantaged areas (1st quintile) | 0.19 | 0.39 | 0.00 | 1.00 |
| Lives in next most disadvantaged areas (2nd quintile) | 0.21 | 0.41 | 0.00 | 1.00 |
| Lives in next most advantaged areas (4th quintile) | 0.21 | 0.41 | 0.00 | 1.00 |
| Lives in the most advantaged areas (5th quintile) | 0.19 | 0.39 | 0.00 | 1.00 |
| Lives in a non-capital city | 0.33 | 0.47 | 0.00 | 1.00 |
| Employed | 0.62 | 0.49 | 0.00 | 1.00 |
| Household income | 674.36 | 545.28 | -8.86 | 5668.75 |
| Too much fuss made about COVID-19 | 0.20 | 0.40 | 0.00 | 1.00 |
| Social distancing behaviour | -0.04 | 1.02 | -2.46 | 1.71 |
| Downloaded the COVID-Safe App | 0.46 | 0.50 | 0.00 | 1.00 |
| Voting intention – Labour | 0.32 | 0.47 | 0.00 | 1.00 |
| Voting intention – Greens | 0.12 | 0.32 | 0.00 | 1.00 |
| Voting intention – Other | 0.06 | 0.24 | 0.00 | 1.00 |
| Voting intention – Don’t know | 0.04 | 0.20 | 0.00 | 1.00 |
| Confident in state or territory government | 0.71 | 0.45 | 0.00 | 1.00 |
| Confident in GPs | 0.87 | 0.34 | 0.00 | 1.00 |
| Support for migration | 6.55 | 2.44 | 0.00 | 10.00 |
| Populism | 18.76 | 4.22 | 6.00 | 30.00 |
| Religiosity | 3.44 | 3.12 | 0.00 | 10.00 |

Source:	ANUpoll, April, May and August 2020.

Notes:	Descriptive statistics from Ordered probit model. N = 2,261.

**Table A.3	Correlates of vaccine resistance and hesitancy – Demographics only, ordered probit**

| Explanatory variables | Vaccine likelihood | |
| --- | --- | --- |
|  | Coeff | Signif. |
| Victoria | 0.031 |  |
| Female | -0.109 | * |
| Aged 18 to 24 years | 0.135 |  |
| Aged 25 to 34 years | -0.052 |  |
| Aged 45 to 54 years | -0.063 |  |
| Aged 55 to 64 years | 0.233 | ** |
| Aged 65 to 74 years | 0.363 | *** |
| Aged 75 years plus | 0.538 | *** |
| Indigenous | -0.033 |  |
| Born overseas in a main English speaking country | -0.074 |  |
| Born overseas in a non-English speaking country | -0.028 |  |
| Speaks a language other than English at home | -0.124 |  |
| Has not completed Year 12 or post-school qualification | -0.079 |  |
| Has a post graduate degree | 0.278 | ** |
| Has an undergraduate degree | 0.206 | ** |
| Has a Certificate III/IV, Diploma or Associate Degree | -0.012 |  |
| Lives in the most disadvantaged areas (1st quintile) | -0.189 | * |
| Lives in next most disadvantaged areas (2nd quintile) | -0.016 |  |
| Lives in next most advantaged areas (4th quintile) | -0.172 | * |
| Lives in the most advantaged areas (5th quintile) | -0.020 |  |
| Lives in a non-capital city | -0.073 |  |
| Employed | -0.005 |  |
| Household income | 0.000 | *** |
| Cut1 | -1.506 |  |
| Cut2 | -1.056 |  |
| Cut3 | -0.097 |  |

Source:	ANUpoll, April, May and August 2020.

Notes:	Ordered probit model. N = 2,717. Coefficients that are statistically significant at the 1 per cent level of significance are labelled ***; those significant at the 5 per cent level of significance are labelled **, and those significant at the 10 per cent level of significance are labelled *.

**Table A.4	Correlates of vaccine resistance and hesitancy – Demographics plus health variables, ordered probit**

| Explanatory variables | Vaccine likelihood | |
| --- | --- | --- |
|  | Coeff | Signif. |
| Victoria | 0.030 |  |
| Female | -0.130 | ** |
| Aged 18 to 24 years | 0.176 |  |
| Aged 25 to 34 years | -0.045 |  |
| Aged 45 to 54 years | -0.073 |  |
| Aged 55 to 64 years | 0.232 | ** |
| Aged 65 to 74 years | 0.438 | *** |
| Aged 75 years plus | 0.563 | *** |
| Indigenous | -0.071 |  |
| Born overseas in a main English speaking country | -0.056 |  |
| Born overseas in a non-English speaking country | -0.052 |  |
| Speaks a language other than English at home | -0.113 |  |
| Has not completed Year 12 or post-school qualification | -0.104 |  |
| Has a post graduate degree | 0.250 | ** |
| Has an undergraduate degree | 0.177 | * |
| Has a Certificate III/IV, Diploma or Associate Degree | -0.057 |  |
| Lives in the most disadvantaged areas (1st quintile) | -0.220 | ** |
| Lives in next most disadvantaged areas (2nd quintile) | -0.030 |  |
| Lives in next most advantaged areas (4th quintile) | -0.171 |  |
| Lives in the most advantaged areas (5th quintile) | -0.016 |  |
| Lives in a non-capital city | -0.083 |  |
| Employed | 0.050 |  |
| Household income | 0.000 | *** |
| Self rated health - good | 0.087 |  |
| Impact of disability or chronic illness on activities of daily living - a lot | 0.078 |  |
| Impact of disability or chronic illness on activities of daily living - a little | -0.038 |  |
| Cut1 | -1.487 |  |
| Cut2 | -1.043 |  |
| Cut3 | -0.074 |  |

Source:	ANUpoll, April, May and August 2020.

Notes:	Ordered probit model. N = 2,510 . Coefficients that are statistically significant at the 1 per cent level of significance are labelled ***; those significant at the 5 per cent level of significance are labelled **, and those significant at the 10 per cent level of significance are labelled *.

**Table A.5	Correlates of vaccine resistance and hesitancy – Demographic plus COVID-19 variables, ordered probit**

| Explanatory variables | Vaccine likelihood | |
| --- | --- | --- |
|  | Coeff | Signif. |
| Victoria | -0.192 | * |
| Female | -0.156 | ** |
| Aged 18 to 24 years | 0.270 |  |
| Aged 25 to 34 years | -0.030 |  |
| Aged 45 to 54 years | -0.078 |  |
| Aged 55 to 64 years | 0.259 | ** |
| Aged 65 to 74 years | 0.359 | *** |
| Aged 75 years plus | 0.536 | *** |
| Indigenous | 0.076 |  |
| Born overseas in a main English speaking country | -0.072 |  |
| Born overseas in a non-English speaking country | -0.011 |  |
| Speaks a language other than English at home | -0.163 |  |
| Has not completed Year 12 or post-school qualification | -0.116 |  |
| Has a post graduate degree | 0.156 |  |
| Has an undergraduate degree | 0.172 |  |
| Has a Certificate III/IV, Diploma or Associate Degree | -0.085 |  |
| Lives in the most disadvantaged areas (1st quintile) | -0.160 |  |
| Lives in next most disadvantaged areas (2nd quintile) | 0.006 |  |
| Lives in next most advantaged areas (4th quintile) | -0.129 |  |
| Lives in the most advantaged areas (5th quintile) | -0.025 |  |
| Lives in a non-capital city | -0.033 |  |
| Employed | 0.019 |  |
| Household income | 0.000 | *** |
| Worried about yourself or family or friends contracting COVID-19 | 0.104 |  |
| Tested for COVID-19 | 0.077 |  |
| Downloaded the COVID-Safe App | 0.333 | *** |
| Too much fuss made about COVID-19 | -0.395 | *** |
| Social distancing behaviour | 0.151 | *** |
| Cut1 | -1.545 |  |
| Cut2 | -1.089 |  |
| Cut3 | -0.078 |  |

Source:	ANUpoll, April, May and August 2020.

Notes:	Ordered probit model. N = 2,374. Coefficients that are statistically significant at the 1 per cent level of significance are labelled ***; those significant at the 5 per cent level of significance are labelled **, and those significant at the 10 per cent level of significance are labelled *.

**Table A.6	Correlates of vaccine resistance and hesitancy – Demographics plus political and social attitudes and trust variables, ordered probit**

| Explanatory variables | Vaccine likelihood | |
| --- | --- | --- |
| Victoria | 0.056 |  |
| Female | -0.129 | * |
| Aged 18 to 24 years | 0.179 |  |
| Aged 25 to 34 years | -0.024 |  |
| Aged 45 to 54 years | -0.009 |  |
| Aged 55 to 64 years | 0.294 | *** |
| Aged 65 to 74 years | 0.432 | *** |
| Aged 75 years plus | 0.626 | *** |
| Indigenous | 0.136 |  |
| Born overseas in a main English speaking country | -0.125 |  |
| Born overseas in a non-English speaking country | -0.041 |  |
| Speaks a language other than English at home | -0.099 |  |
| Has not completed Year 12 or post-school qualification | -0.079 |  |
| Has a post graduate degree | 0.243 | * |
| Has an undergraduate degree | 0.177 | * |
| Has a Certificate III/IV, Diploma or Associate Degree | -0.031 |  |
| Lives in the most disadvantaged areas (1st quintile) | -0.223 | ** |
| Lives in next most disadvantaged areas (2nd quintile) | -0.043 |  |
| Lives in next most advantaged areas (4th quintile) | -0.183 | * |
| Lives in the most advantaged areas (5th quintile) | -0.050 |  |
| Lives in a non-capital city | -0.072 |  |
| Employed | 0.016 |  |
| Household income | 0.000 | *** |
| Voting intention – Labour | 0.162 | * |
| Voting intention – Greens | -0.003 |  |
| Voting intention – Other | -0.028 |  |
| Voting intention – Don’t know | -0.117 |  |
| Confident in Federal Government | -0.122 |  |
| Confident in state or territory government | 0.280 | *** |
| Confident in GPs | 0.328 | *** |
| Altruism | -0.007 |  |
| Support for migration | 0.029 | * |
| Religiosity | -0.018 | * |
| Social trust | -0.008 |  |
| Populism | -0.031 | *** |
| Authoritarianism | -0.010 |  |
| Cut1 | -1.639 |  |
| Cut2 | -1.178 |  |
| Cut3 | -0.190 |  |

Source:	ANUpoll, April, May and August 2020.

Notes:	Ordered probit model. N = 2,359. Coefficients that are statistically significant at the 1 per cent level of significance are labelled ***; those significant at the 5 per cent level of significance are labelled **, and those significant at the 10 per cent level of significance are labelled *.

**Table A.7	Correlates of vaccine resistance and hesitancy – Final model, ordered probit**

| Explanatory variables | Vaccine likelihood | |
| --- | --- | --- |
|  | Coeff | Signif. |
| Victoria | -0.200 | * |
| Female | -0.153 | ** |
| Aged 18 to 24 years | 0.213 |  |
| Aged 25 to 34 years | -0.004 |  |
| Aged 45 to 54 years | -0.033 |  |
| Aged 55 to 64 years | 0.279 | ** |
| Aged 65 to 74 years | 0.391 | *** |
| Aged 75 years plus | 0.565 | *** |
| Indigenous | 0.080 |  |
| Born overseas in a main English speaking country | -0.127 |  |
| Born overseas in a non-English speaking country | -0.003 |  |
| Speaks a language other than English at home | -0.132 |  |
| Has not completed Year 12 or post-school qualification | -0.096 |  |
| Has a post graduate degree | 0.190 |  |
| Has an undergraduate degree | 0.154 |  |
| Has a Certificate III/IV, Diploma or Associate Degree | -0.100 |  |
| Lives in the most disadvantaged areas (1st quintile) | -0.212 | * |
| Lives in next most disadvantaged areas (2nd quintile) | -0.005 |  |
| Lives in next most advantaged areas (4th quintile) | -0.166 |  |
| Lives in the most advantaged areas (5th quintile) | -0.073 |  |
| Lives in a non-capital city | -0.045 |  |
| Employed | 0.028 |  |
| Household income | 0.000 | *** |
| Too much fuss made about COVID-19 | -0.346 | *** |
| Social distancing behaviour | 0.172 | *** |
| Downloaded the COVID-Safe App | 0.282 | *** |
| Voting intention – Labour | 0.190 | ** |
| Voting intention – Greens | 0.034 |  |
| Voting intention – Other | 0.169 |  |
| Voting intention – Don’t know | -0.033 |  |
| Confident in state or territory government | 0.190 | ** |
| Confident in GPs | 0.270 | *** |
| Support for migration | 0.023 |  |
| Populism | -0.022 | ** |
| Religiosity | -0.022 | * |
| Cut1 | -1.563 |  |
| Cut2 | -1.090 |  |
| Cut3 | -0.054 |  |

Source:	ANUpoll, April, May and August 2020.

Notes:	Ordered probit model. N = 2,261. Coefficients that are statistically significant at the 1 per cent level of significance are labelled ***; those significant at the 5 per cent level of significance are labelled **, and those significant at the 10 per cent level of significance are labelled *.
